# Characterizing the use of the ICD-10 Code for Long COVID in 3 US Healthcare Systems

**DOI:** 10.1101/2023.02.12.23285701

**Authors:** Harrison G Zhang, Jacqueline P Honerlaw, Monika Maripuri, Malarkodi Jebathilagam Samayamuthu, Brendin R Beaulieu-Jones, Huma S Baig, Sehi L’Yi, Yuk-Lam Ho, Michele Morris, Vidul Ayakulangara Panickan, Xuan Wang, Chuan Hong, Griffin M Weber, Katherine P Liao, Shyam Visweswaran, Bryce W.Q. Tan, William Yuan, Nils Gehlenborg, Sumitra Muralidhar, Rachel B Ramoni, The Consortium for Clinical Characterization of COVID-19 by EHR (4CE), Isaac S Kohane, Zongqi Xia, Kelly Cho, Tianxi Cai, Gabriel A Brat

**Author notes:** Corresponding author: Gabriel A Brat, MD, MPH. Department of Biomedical Informatics, Harvard Medical School, Boston, MA, United States. These authors contributed equally (Isaac S Kohane, Zongqi Xia, Kelly Cho, Tianxi Cai, Gabriel A Brat).

## Abstract

The International Classification of Diseases (ICD)-10 code (U09.9) for post-acute sequelae of COVID-19 (PASC) was introduced in October of 2021. As researchers seek to leverage this billing code for research purposes in large scale real-world studies of PASC, it is of utmost importance to understand the functional use of the code by healthcare providers and the clinical characteristics of patients who have been assigned this code. To this end, we operationalized clinical case definitions of PASC using World Health Organization and Centers for Disease Control guidelines. We then chart reviewed 300 patients with COVID-19 from three participating healthcare systems of the 4CE Consortium who were assigned the U09.9 code. Chart review results showed the average positive predictive value (PPV) of the U09.9 code ranged from 40.2% to 65.4% depending on which definition of PASC was used in the evaluation. The PPV of the U09.9 code also fluctuated significantly between calendar time periods. We demonstrated the potential utility of textual data extracted from natural language processing techniques to more comprehensively capture symptoms associated with PASC from electronic health records data. Finally, we investigated the utilization of long COVID clinics in the cohort of patients. We observed that only an average of 24.0% of patients with the U09.9 code visited a long COVID clinic. Among patients who met the WHO PASC definition, only an average of 35.6% visited a long COVID clinic.

## INTRODUCTION

As the significant clinical, public health, and economic ramifications of the post-acute sequelae of COVID-19 (PASC) become increasingly evident, the biomedical enterprise has raced to better understand and functionally characterize the PASC that are estimated to effect 10% to 40% of all people infected with SARS-CoV-2.^1–4^ Real-world data such as electronic health records (EHR) data has the potential to capture the wide spectrum of clinical features attributed to PASC in diverse patient populations.^5^

To this end, the International Classification of Diseases (ICD)-10 codes is a system used by healthcare providers to code all diagnoses for claims processing and is also commonly used for EHR research purposes to capture outcomes and define cohorts in clinical studies.^6^ The World Health Organization (WHO) introduced an ICD-10 code specifically for long COVID (U09.9) in October of 2021.^7^ This code can enable retrospective studies of long COVID using EHR data by helping researchers define clinical cohorts and ascertain outcomes. Given the very recent rollout of the code and the inherent ambiguity in PASC clinical case definitions, it is therefore of great importance to better understand how the new ICD-10 code for long COVID is being used in real-world settings. In this study, we chart reviewed patients with COVID-19 who were assigned the U09.9 code to understand how well these patients met clinical case definitions of PASC that we operationalized according to global health agency guidelines. We further assessed how well various EHR data types captured the constellation of symptoms associated with PASC and how well the U09.9 code predicted chart review-derived labels of PASC.

## METHODS

To investigate the clinical characteristics of patients who are assigned this code in real-world settings, we convened a group of researchers to perform manual medical record review of COVID-19 patients coded with U09.9 identified at three U.S. healthcare systems: the Beth Israel Deaconess Medical Center, University of Pittsburgh Medical Center, and US Veterans Health Administration. These three health systems collectively serve over 15 million patients each year. We collected detailed information pertaining to PASC using a randomly sampled group of 100 patients at each healthcare system (300 total) with at least one ICD-10 U09.9 for PASC and a documented infection with SARS-CoV-2. A SARS-CoV-2 infection was defined as an positive polymerase chain reaction test result for SARS-CoV-2 or an ICD-10 U07.1 code for COVID-19 prior to the ICD-10 code for PASC.

Currently, various clinical definitions of PASC exist, and they encompass a range of symptoms pertaining to several organ systems. Common PASC symptoms such as headache, fatigue, and persistent pain are also present across a myriad of other conditions, making it an inherently difficult syndrome to diagnose. To evaluate the positive predictive value (PPV) of the ICD-10 code for predicting PASC, we operationalized the WHO and United States Centers for Disease Control’s (CDC) clinical case definitions of PASC after reviewing published guidelines.^8,9^ A robust chart review procedure for PASC was also developed and implemented at the three healthcare systems. The chart review procedure determined true disease status for PASC as defined using WHO and CDC guidelines and other relevant clinical characteristics of the patients. These clinical characteristics include: hospitalization status, long COVID clinical visits, new-onset symptoms after SARS-CoV-2 infection, and COVID-19 testing modality.

We further evaluated the positive predictive value (PPV) and negative predictive value (NPV) of the U09.9 code in predicting a patient’s true disease status at the US Veterans Health Administration. These evaluation results are stratified into two periods based on their initial infection date. Patients were assigned to the first period if they had an initial infection date before September 1, 2021 and the second period if they had an initial infection date on or after September 1, 2021.

We hypothesized that existing administrative coding schemas such as the ICD-10 may not be well-positioned to comprehensively capture PASC symptoms (such as the ICD-10 system). This is because PASC appears to be a clinical syndrome that encompasses a constellation of sometimes non-specific symptoms (*e*.*g*., pain, fatigue, brain fog) that are not well represented or not often coded using administrative coding schemas.^5,10^ We investigated whether or not textual data extracted using natural language processing pipelines can better capture the symptomatology associated with PASC for a patient that was extracted in chart review. For seven common symptoms associated with PASC, we examined the data capture of each symptom by (1) ICD-10 codes alone, (2) natural language processing (NLP) of clinical narratives (*e*.*g*. clinician notes and discharge summaries) alone, (3) both, or (4) either codes or NLP.^5,8,9,11–14^ Diagnosis codes and concept unique identifiers (CUIs) that were mapped to each of the 7 PASC symptoms are listed in Supplementary Table 6.

## RESULTS

We used guidelines published by the WHO and CDC to operationalize robust definitions of PASC used in the chart review. Specifically, we operationalized the WHO’s clinical case definition of PASC to require that a patient present with at least two new-onset symptoms that each persisted for at least 60 days after initial infection. Additionally, we considered an alternative WHO clinical case definition of PASC which only requires a single new-onset symptom that persisted for at least 60 days. We further operationalized the CDC’s clinical case definition of PASC to require that a patient present with at least one new-onset symptom that persisted for at least 30 days. We did not include exacerbations of existing disease to improve consistency for this exercise as it is often difficult to identify exacerbations in the EHR. Further details of the chart review procedure and the clinical case definitions are provided in the supplementary materials.

Chart review was performed on a total of 300 patients infected with COVID-19 who were assigned the U09.9 code across three US healthcare systems. Pertinent clinical characteristics of these patients are summarized in Figure 1. Among patients coded with U09.9, the PPV of the U09.9 code in predicting true disease status defined from chart review was 40.2% for the WHO definition, 58.3% for the single symptom WHO definition, and 65.4% for the CDC definition (Figure 1A). The observed pattern in the PPV of the U09.9 code across definitions was that the stricter definitions, such as the WHO one, yielded lower PPVs. We further report the PPV and NPV of the U09.9 code stratified by calendar time period of the patient’s initial infection date (Table 1). We observed a similar pattern of the PPV when comparing between the three definitions. For each of the three definitions, the PPV of the code was lower in the cohort of patients with an initial infection date of on or after October 1, 2021 compared to the cohort of patients with an initial infection date of before October 1, 2021. The NPV of the code was highest among PASC definitions with stricter criteria. However in contrast to the PPV, the NPV of the code stayed relatively constant for a given PASC definition between calendar time periods.

**Table 1:** Positive predictive value (PPV) and Negative Predictive Value (NPV) of the U09.9 Code at the US Veterans Health Administration.

| PASC Definitions | Calendar Time Period | PPV (95% CI) | NPV (95% CI) |
| --- | --- | --- | --- |
| WHO | Before October 1, 2021 | 0.474 (0.373-0.574) | 0.927 (0.892-0.962) |
| WHO | On or after October 1, 2021 | 0.229 (0.167-0.291) | 0.917 (0.872-0.962) |
| WHO1 | Before October 1, 2021 | 0.705 (0.614-0.797) | 0.817 (0.749-0.884) |
| WHO1 | On or after October 1, 2021 | 0.436 (0.363-0.508) | 0.861 (0.781-0.941) |
| CDC | Before October 1, 2021 | 0.747 (0.660-0.835) | 0.716 (0.629-0.802) |
| CDC | On or after October 1, 2021 | 0.603 (0.532-0.675) | 0.750 (0.623-0.877) |

**Figure 1:**
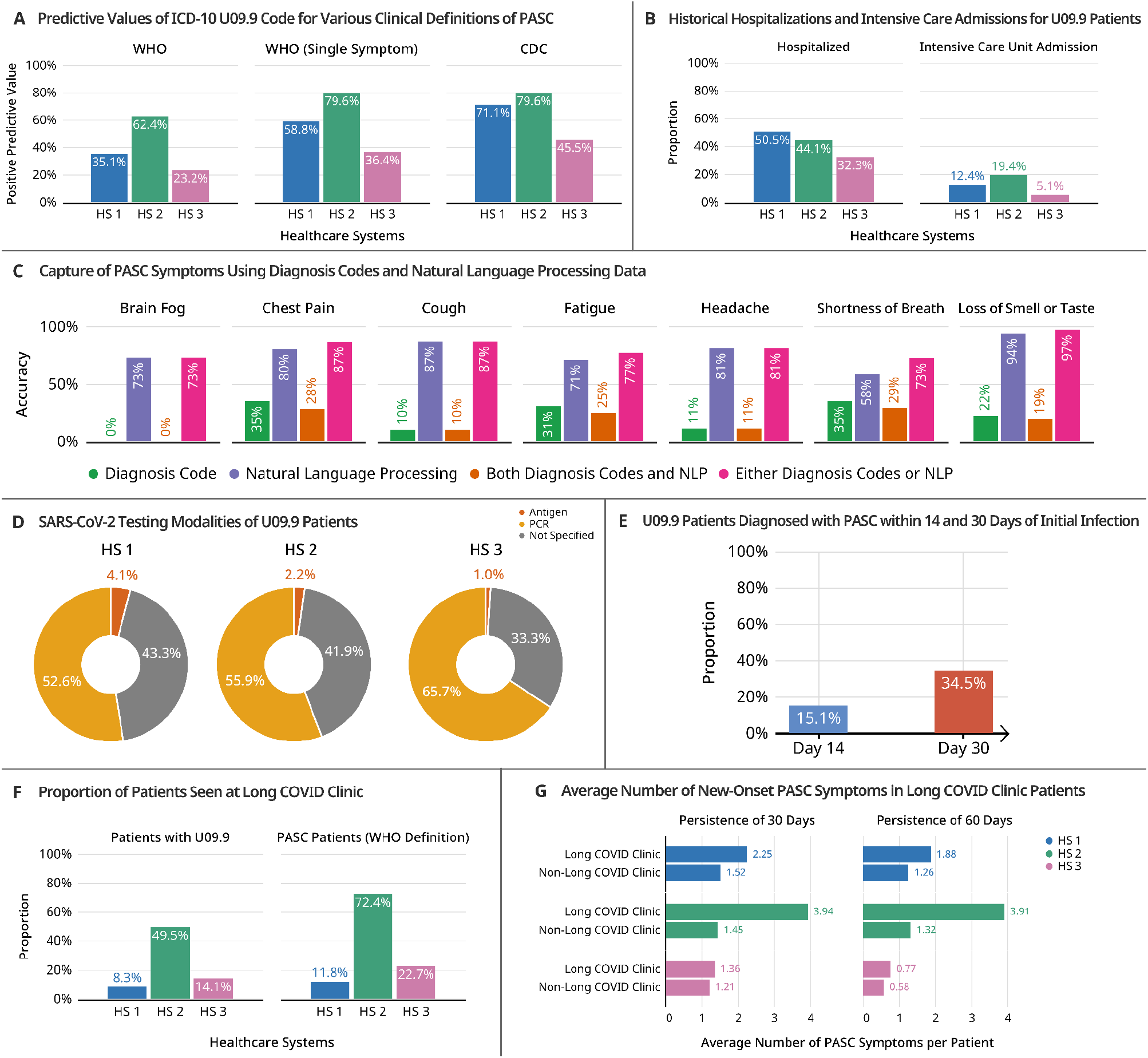
Clinical characteristics of patients with the PASC code (ICD-10 U09.9) in three healthcare systems (HS). (A) Predictive value of PASC code for clinical definitions of PASC. (B) Hospitalizations and admissions to intensive care units. (C) Data capture of PASC symptoms abstracted from chart review using diagnostic codes, textual data from natural language processing, both data elements, and either data elements. (D) Testing modalities captured in the electronic health records data. (E) Proportion of patients diagnosed within 14 and 30 days from initial infection. (F) Long COVID clinic use among patients with the PASC code and among patients who met WHO PASC clinical definitions. (G) Comparison of the average number of PASC symptoms per patient in patients who visited a long COVID clinic and among those who never visited a long COVID clinic.

For seven common symptoms associated with PASC, we report the ability of ICD-10 codes and textual data to capture these symptoms in patients (Figure 1C). Across all the symptoms considered, using only textual data elements as extracted with NLP better captured symptoms identified through chart review compared to using ICD-10 codes alone. The improvement in data capture when using textual data was most evident for the neurologic symptoms that were considered, which include brain fog, headaches, and loss of smell and taste (Figure 1C). Further, we observed the most complete capture of all symptoms with EHR data elements when using either textual data or ICD-10 codes.

Among the sampled patients, a SARS-CoV-2 infection was most commonly documented in the patients’ EHR with a PCR test result (Figure 1D). We observed that rapid antigen testing is less commonly documented in the EHR and accounted for a very small portion of documented infections in the patient sample (Figure 1D). Furthermore, we were not able to identify the COVID-19 testing modality for at least a third of the patients using EHR data elements (Figure 1D). The unstable nature of infection dates ascertained from EHR data may explain the puzzling observation that an average of 34.5% of patients had a U09.9 code within the first 30 days of an index date based on the first positive SARS-CoV-2 test abstracted from EHR data elements (Figure 1E).

Finally, we investigated the use of long COVID clinics by patients in our cohort. We observed that only an average of 24.0% of patients with the U09.9 code visited a long COVID clinic (Figure 1F). Among patients who met the WHO PASC definition, only an average of 35.6% visited a long COVID clinic (Figure 1F). Furthermore, across all three healthcare systems, patients who visited a long COVID clinic were on average diagnosed with more new-onset symptoms compared to patients who never visited a long COVID clinic (Figure 1G).

Future studies of PASC which utilize large-scale real-world data and the ICD-10 diagnosis code for PASC must be robustly designed to account for the limitations of the code identified in this study that have the potential to bias findings.

## Data Availability

Summary level data produced in the present study are available upon request to the authors.

