## supplementary materials for "Characterizing the use of the ICD-10 Code for Long COVID in 3 US Healthcare Systems"

### Chart Review Procedure for Post-Acute Sequelae of COVID-19 (PASC)

#### Definition of Long COVID-19

The definition of long covid for this chart review is drawn from the World Health Organization (WHO) Consensus Definition 2021:

> *"Post COVID-19 condition occurs in individuals with **a history of probable or confirmed SARS-CoV-2** infection, **usually 3 months from the onset of COVID-19 with symptoms that last for at least 2 months and cannot be explained by an alternative diagnosis.** Common symptoms include **fatigue, shortness of breath, cognitive dysfunction** but also others (see **Table 3** and **Annex 2**) which generally have an **impact on everyday functioning.** Symptoms may be **new onset,** following initial recovery from an acute COVID- 19 episode, or **persist** from the initial illness. Symptoms may also **fluctuate** or **relapse** over time. A separate definition may be applicable for children."*

The following operational definition was adapted from WHO to classify a patient with long COVID-19 for this chart review:

1. Symptom/pathology must be new onset, occurring after COVID-19 diagnosis.
2. Symptom/pathology must not be an exacerbation of an existing objective pre-COVID pathology.
3. Symptom/pathology must persist for at least 2 months (60 days).(* 1 month by CDC definition.)
4. Symptoms must start within 6 months after initial COVID infection.
5. Review charts of patients who have atleast 3 months of data post COVID-19 infection.
6. The patient chart must include at least 1 symptom/pathology from the Core set. *(See Table 5 for Core symptom list)*
7. IF only 1 Core symptom/pathology is present, the patient chart must also include at least 1 symptom/disease from Extended symptom cluster set. *(See Table 5 for Extended symptom cluster list)*
   Fluctuating symptoms can be counted if the symptoms are present for the majority of the 60 day period.

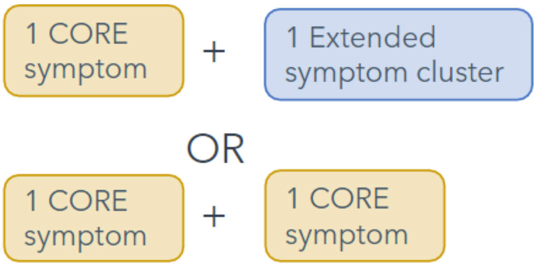


A second definition for long COVID-19 was adapted from the Centers for Disease Control and Prevention (CDC). The CDC definition uses a 1 month duration for symptoms and the items marked in "*" denote differences from the WHO definition.

1. Symptom/pathology must be new onset, occurring after COVID-19 diagnosis.
2. Symptom/pathology must not be an exacerbation of an existing objective pre-COVID pathology.
3. Symptom/pathology must persist for at least 1 month.*
4. Symptoms must start within 6 months after initial COVID infection.

5. Review charts of patients who have atleast 3 months of data post COVID-19 infection.
6. The patient chart must include at least 1 symptom/pathology from the Core set. *(See Table 5 for Core symptom list)*
7. IF only 1 Core symptom/pathology is present, the patient chart must also include at least 1 symptom/disease from Extended symptom cluster set. *(See Table 5 for Extended symptom cluster list)*
8. Fluctuating symptoms can be counted if the symptoms are present for the majority of the 30 day period.*

##### Supplementary Table 1: Chart Sampling

| | |
|---|---|
| | • COVID Positive (PCR positive test or U07.1 ICD Code)<br>• Presence of ICD-10 code U09.9 |

10% overlap in chart reviews was coordinated between reviewers in order to ensure replicability and consistency in the process and across institutions. Any discrepancies were resolved by shared decision making and consensus.

##### Supplementary Table 2: Data Sources for Chart Review

| | |
|---|---|
| **ICD-10 Codes** | All available ICD codes and dates (including U09.9) |
| **Labs** | List of all positive COVID-19 tests and date |
| **Patient Notes** | All notes available in data pull timeframe |
| **COVID-19 related hospitalizations** | Admission code, date, discharge date |

#### Reviewer Classifications

##### Supplementary Table 3: COVID Diagnosis and Baseline Conditions

| ***Field*** | ***Description*** |
|---|---|
| First Positive COVID Test | Date of first positive COVID-19 lab test. This may be found in the structured lab data or in free text (if performed outside of health system) |
| Location | Location of COVID-19 diagnosis (within the system or at an outside location) |

##### Supplementary Table 4: COVID Vaccination

| ***Field*** | ***Description*** |
|---|---|
| Vaccination status | Was the patient ever vaccinated? |
| Vaccination date | Date of first vaccination |
| Location | Was the patient vaccinated in the hospital system or at an outside location |

##### Supplementary Table 5: COVID-19 Related Symptom(s) and Duration

| ***Field*** | ***Description*** | ***Data entered*** |
|---|---|---|
| Core Symptom Cluster | Specific, very common symptoms that are nearly always present. | **Check all that apply**<br>Loss/Changes of smell/taste<br>Abdominal pain |

<table>
<tr>
<td></td>
<td>Only capture:
<ul>
<li>Symptoms with onset after COVID-19 infection</li>
<li>Symptoms that start within 6 months of covid infection</li>
<li>Symptoms that persist for at least 30 days (CDC Definition)</li>
<li>Symptoms that persist for at least 60 days (WHO Definition)</li>
</ul></td>
<td><table>
<tr><td>Diarrhea</td></tr>
<tr><td>Headache</td></tr>
<tr><td>Persistent cough</td></tr>
<tr><td>Fatigue</td></tr>
<tr><td>Fever</td></tr>
<tr><td>Shortness of breath</td></tr>
<tr><td>Chest pain</td></tr>
<tr><td>Unusual muscle pains</td></tr>
<tr><td>Cognitive dysfunction/confusion/”fog”</td></tr>
</table></td>
</tr>
<tr>
<td>Extended Symptom Cluster</td>
<td>Manual grouping of symptoms for ease of chart review. Based on phecode groups.<br><br>Only capture:
<ul>
<li>Symptoms with onset after COVID-19 infection</li>
<li>Symptoms that start within 6 months of covid infection</li>
<li>Symptoms that persist for at least 30 days (CDC Definition)</li>
<li>Symptoms that persist for at least 60 days (WHO Definition)</li>
</ul>
Start date is for the first noted symptom in the cluster</td>
<td><table>
<tr><th>Check all that apply</th><th>Symptom(s) **<br>(free text)</th></tr>
<tr><td>Cardiac/Circulatory</td><td></td></tr>
<tr><td>Dermatologic</td><td></td></tr>
<tr><td>Endocrine/metabolic</td><td></td></tr>
<tr><td>Gastrointestinal</td><td></td></tr>
<tr><td>Genitourinary</td><td></td></tr>
<tr><td>Hematopoietic</td><td></td></tr>
<tr><td>Infectious disease</td><td></td></tr>
<tr><td>Mental disorders</td><td></td></tr>
<tr><td>Musculoskeletal</td><td></td></tr>
<tr><td>Neurologic</td><td></td></tr>
<tr><td>Respiratory</td><td></td></tr>
<tr><td>Sense organs</td><td></td></tr>
<tr><td>Symptoms</td><td></td></tr>
</table>
**List each of the symptoms identified in the cluster</td>
</tr>
<tr>
<td>Long COVID</td>
<td>Does that patient meet the definition of long COVID as described using either the WHO or CDC guidelines?</td>
<td><ul>
<li>At least 1 symptom from the Core Symptom Cluster</li>
<li>IF only 1 Core symptom/pathology present, must include at least 1 symptom/disease from Extended set.</li>
</ul></td>
</tr>
</table>

#### Supplementary Figure 1: Timeline of defining Long COVID and capturing symptoms from COVID infection date

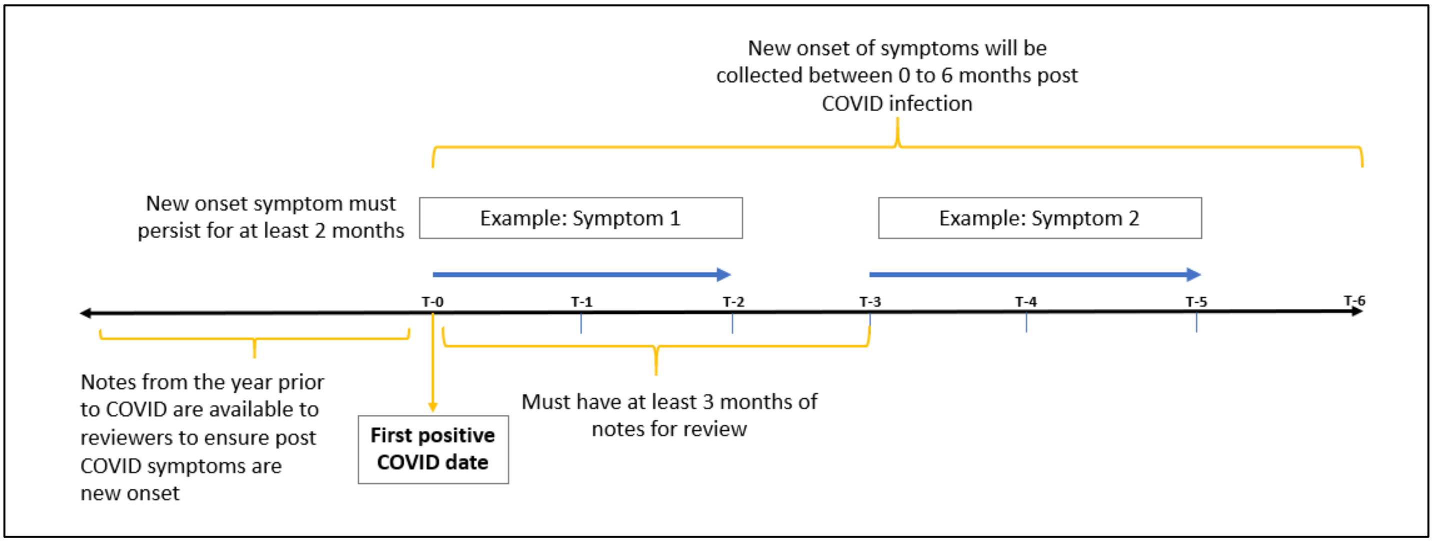

Supplementary Table 6: COVID-19 Related Symptom(s) and Duration

| CORE_SYMPTOM | CONCEPT_TYPE | CONCEPT_CODE |
|---|---|---|
| Loss/Changes of smell/taste | PheCode | 350.6 |
| Abdominal pain | PheCode | 785 |
| Diarrhea | PheCode | 561.1 |
| Headache | PheCode | 306.9 |
| Headache | PheCode | 339 |
| Cough | PheCode | 512.8 |
| Fatigue | PheCode | 798.1 |
| Fatigue | PheCode | 798 |
| Fever | PheCode | 783 |
| Shortness of breath | PheCode | 512.7 |
| Chest pain | PheCode | 418 |
| Unusual muscle pains | PheCode | 772 |
| Cognitive dysfunction/confusion/,Äùfog,Äù | PheCode | NA |
| Loss/Changes of smell/taste | CUI | C2364082 |
| Loss/Changes of smell/taste | CUI | C1510410 |
| Loss/Changes of smell/taste | CUI | C0481703 |
| Loss/Changes of smell/taste | CUI | C0234259 |
| Loss/Changes of smell/taste | CUI | C0423570 |
| Loss/Changes of smell/taste | CUI | C0240327 |
| Loss/Changes of smell/taste | CUI | C0039338 |
| Loss/Changes of smell/taste | CUI | C0481703 |
| Loss/Changes of smell/taste | CUI | C0578994 |
| Loss/Changes of smell/taste | CUI | C0423564 |
| Loss/Changes of smell/taste | CUI | C0003126 |
| Loss/Changes of smell/taste | CUI | C2364111 |
| Abdominal pain | CUI | C0000737 |
| Abdominal pain | CUI | C2585575 |
| Abdominal pain | CUI | C0232491 |
| Abdominal pain | CUI | C0232495 |
| Abdominal pain | CUI | C0232492 |
| Abdominal pain | CUI | C0589386 |
| Abdominal pain | CUI | C0476289 |
| Abdominal pain | CUI | C0344304 |
| Abdominal pain | CUI | C0522061 |
| Abdominal pain | CUI | C1282002 |
| Abdominal pain | CUI | C0581869 |

| | | |
|---|---|---|
| Abdominal pain | CUI | C2586208 |
| Abdominal pain | CUI | C0423644 |
| Abdominal pain | CUI | C4047369 |
| Abdominal pain | CUI | C0563276 |
| Abdominal pain | CUI | C0563277 |
| Diarrhea | CUI | C0011991 |
| Diarrhea | CUI | C0401151 |
| Diarrhea | CUI | C0474496 |
| Diarrhea | CUI | C0740441 |
| Diarrhea | CUI | C0151594 |
| Diarrhea | CUI | C1443924 |
| Diarrhea | CUI | C0521604 |
| Diarrhea | CUI | C0232711 |
| Diarrhea | CUI | C0156173 |
| Diarrhea | CUI | C0474495 |
| Diarrhea | CUI | C0267436 |
| Diarrhea | CUI | C0152522 |
| Diarrhea | CUI | C0267664 |
| Headache | CUI | C0018681 |
| Headache | CUI | C0009088 |
| Headache | CUI | C0033893 |
| Headache | CUI | C0393738 |
| Headache | CUI | C0752147 |
| Headache | CUI | C0751185 |
| Headache | CUI | C0032816 |
| Headache | CUI | C1168188 |
| Headache | CUI | C0239886 |
| Headache | CUI | C0521668 |
| Headache | CUI | C0231613 |
| Headache | CUI | C0423618 |
| Headache | CUI | C0948396 |
| Headache | CUI | C0744633 |
| Headache | CUI | C0393737 |
| Headache | CUI | C0338486 |
| Headache | CUI | C0423621 |
| Headache | CUI | C0037195 |
| Headache | CUI | C0581880 |
| Headache | CUI | C0393739 |
| Headache | CUI | C0393736 |

| | | |
|---|---|---|
| Headache | CUI | C3661947 |
| Headache | CUI | C0751191 |
| Headache | CUI | C0042376 |
| Headache | CUI | C0474366 |
| Headache | CUI | C2349426 |
| Headache | CUI | C2316225 |
| Headache | CUI | C3853085 |
| Headache | CUI | C0522253 |
| Headache | CUI | C2875239 |
| Headache | CUI | C0393745 |
| Headache | CUI | C0423623 |
| Headache | CUI | C2875221 |
| Headache | CUI | C0744644 |
| Headache | CUI | C0581875 |
| Headache | CUI | C0423619 |
| Cough | CUI | C0010200 |
| Cough | CUI | C0010201 |
| Cough | CUI | C0850149 |
| Cough | CUI | C0239134 |
| Cough | CUI | C1277295 |
| Cough | CUI | C0423730 |
| Cough | CUI | C0687152 |
| Cough | CUI | C0562483 |
| Cough | CUI | C0234866 |
| Cough | CUI | C0231912 |
| Cough | CUI | C0231911 |
| Cough | CUI | C0239133 |
| Cough | CUI | C0151574 |
| Cough | CUI | C0455777 |
| Cough | CUI | C0240351 |
| Cough | CUI | C0586750 |
| Cough | CUI | C0574067 |
| Cough | CUI | C4708767 |
| Cough | CUI | C0277870 |
| Cough | CUI | C0694548 |
| Cough | CUI | C0577920 |
| Cough | CUI | C0425497 |
| Cough | CUI | C0566525 |
| Fatigue | CUI | C0015672 |

| | | |
|---|---|---|
| Fatigue | CUI | C0015674 |
| Fatigue | CUI | C0024528 |
| Fatigue | CUI | C2732413 |
| Fever | CUI | C0015967 |
| Fever | CUI | C0424755 |
| Fever | CUI | C0035021 |
| Fever | CUI | C0743841 |
| Fever | CUI | C0239574 |
| Fever | CUI | C1277295 |
| Fever | CUI | C0085594 |
| Fever | CUI | C0424768 |
| Fever | CUI | C0277799 |
| Fever | CUI | C0424749 |
| Fever | CUI | C0424767 |
| Fever | CUI | C0687681 |
| Shortness of breath | CUI | C0235546 |
| Shortness of breath | CUI | C1321587 |
| Shortness of breath | CUI | C0425488 |
| Shortness of breath | CUI | C0423729 |
| Shortness of breath | CUI | C0231898 |
| Shortness of breath | CUI | C0566271 |
| Shortness of breath | CUI | C0574066 |
| Chest pain | CUI | C0008031 |
| Chest pain | CUI | C0238995 |
| Chest pain | CUI | C0522051 |
| Chest pain | CUI | C0232285 |
| Chest pain | CUI | C0476281 |
| Chest pain | CUI | C0008035 |
| Chest pain | CUI | C0476280 |
| Chest pain | CUI | C0423729 |
| Chest pain | CUI | C0262384 |
| Chest pain | CUI | C1740831 |
| Chest pain | CUI | C0232288 |
| Chest pain | CUI | C0232290 |
| Chest pain | CUI | C1299600 |
| Chest pain | CUI | C0232289 |
| Chest pain | CUI | C0392685 |
| Chest pain | CUI | C0029537 |
| Chest pain | CUI | C0541828 |

| | | |
|---|---|---|
| Chest pain | CUI | C0563275 |
| Chest pain | CUI | C0476278 |
| Chest pain | CUI | C0563278 |
| Chest pain | CUI | C0423634 |
| Unusual muscle pains | CUI | C0575064 |
| Unusual muscle pains | CUI | C0242979 |
| Cognitive dysfunction/confusion/Brain fog | CUI | C0009676 |
| Cognitive dysfunction/confusion/ Brain fog | CUI | C0338656 |
| Cognitive dysfunction/confusion/ Brain fog | CUI | C0278061 |
| Cognitive dysfunction/confusion/ Brain fog | CUI | C0233794 |
| Cognitive dysfunction/confusion/ Brain fog | CUI | C0009241 |
| Cognitive dysfunction/confusion/ Brain fog | CUI | C1270972 |
| Cognitive dysfunction/confusion/ Brain fog | CUI | C0701811 |
| Cognitive dysfunction/confusion/ Brain fog | CUI | C0233823 |
| Cognitive dysfunction/confusion/ Brain fog | CUI | C3887551 |
| Cognitive dysfunction/confusion/ Brain fog | CUI | C0752295 |
