## Supplementary material for "Characterizing the use of the ICD-10 Code for Long COVID in 3 US Healthcare Systems": NA

James R Aaron MHA, Giuseppe Agapito PhD, Adem Albayrak, Giuseppe Albi MS, Mario Alessiani MD, FACS, Anna Alloni PhD, Danilo F Amendola MSc, François Angoulvant MD,PhD, Li L.L.J Anthony, Bruce J Aronow PhD, Fatima Ashraf MS, Andrew Atz MD, Paul Avillach MD, PhD, Vidul Ayakulangara Panickan MS, Paula S Azevedo MD, PhD, James Balshi, Ashley Batugo BS, Brett K Beaulieu-Jones PhD, Brendin R Beaulieu-Jones MD, MBA, Douglas S Bell, Antonio Bellasi MD, PhD, Riccardo Bellazzi MS, PhD, Vincent Benoit PhD, Michele Beraghi MS, José Luis Bernal-Sobrino MS, Mélodie Bernaux, Romain Bey, Surbhi Bhatnagar PhD, Alvar Blanco-Martínez MS, Martin Boeker, Clara-Lea Bonzel MSc, John Booth MSc, Silvano Bosari Prof., Florence T Bourgeois MD, MPH, Robert L Bradford, Gabriel A Brat MD, Stéphane Bréant, Nicholas W Brown MEng, Raffaele Bruno MD, William A Bryant PhD, Mauro Bucalo MS, Emily Bucholz MD, PhD, MPH, Anita Burgun, Tianxi Cai ScD, Mario Cannataro M.Sc., Aldo Carmona, Anna Maria Cattelan MD, Charlotte Caucheteux, Julien Champ, Jin Chen PhD, Krista Y Chen BS, Luca Chiovato MD, PhD, Lorenzo Chiudinelli PhD, Kelly Cho PhD, MPH, James J Cimino MD, Tiago K Colicchio PhD, MBA, Sylvie Cormont, Sébastien Cossin, Jean B Craig PhD, Juan Luis Cruz-Bermúdez PhD, Jaime Cruz-Rojo MD, Arianna Dagliati MS, PhD, Mohamad Daniar MSIS, Christel Daniel, Priyam Das PhD, Batsal Devkota, Audrey Dionne MD, Rui Duan PhD, Julien Dubiel, Scott L DuVall PhD, Loic Esteve, Hossein Estiri PhD, Shirley Fan, Robert W Follett BS, Thomas Ganslandt MD, Noelia García-Barrio MS, Lana X Garmire PhD, Nils Gehlenborg, Emily J Getzen MS, Alon Geva MD, MPH, Tomás González González MD, Tobias Gradinger MD, BSc, Alexandre Gramfort, Romain Griffier, Nicolas Griffon, Olivier Grisel, Alba Gutiérrez-Sacristán PhD, pietro h guzzi PhD, Larry Han PhD, David A Hanauer MD, MS, Christian Haverkamp MD, Derek Y Hazard MSc, Bing He PhD, Darren W Henderson BS, Martin Hilka, Yuk-Lam Ho MPH, John H Holmes MS, PhD, Jacqueline P Honerlaw RN, MPH, Chuan Hong PhD, Kenneth M Huling HS, Meghan R Hutch BS, Richard W Issitt DClinP, Anne Sophie Jannot, Vianney Jouhet MD,PhD, Ramakanth Kavuluru PhD, Mark S Keller, Chris J Kennedy PhD, Kate F Kernan MD, Daniel A Key BEng, Katie Kirchoff MSHI, Jeffrey G Klann MEng, PhD, Isaac S Kohane MD, PhD, Ian D Krantz, Detlef Kraska Dr., Ashok K Krishnamurthy PhD, Sehi L'Yi PhD, Trang T Le PhD, Judith Leblanc, Guillaume Lemaitre, Leslie Lenert MD, MS, Damien Leprovost, Molei Liu PhD, Ne Hooi Will Loh MBBS, Qi Long PhD, Sara Lozano-Zahonero PhD, Yuan Luo PhD, Kristine E Lynch PhD, Sadiqa Mahmood, Sarah E Maidlow AA, Adeline Makoudjou MD, Simran Makwana MS, Alberto Malovini PhD, Kenneth D Mandl MD, MPH, Chengsheng Mao PhD, Anupama Maram MS, Monika Maripuri MBBS, MPH, Patricia Martel, Marcelo R Martins MSc, Jayson S Marwaha MD, Aaron J Masino PhD, Maria Mazzitelli MD, PhD, Diego R Mazzotti PhD, Arthur Mensch, Marianna Milano PhD, Marcos F Minicucci MD, PhD, Bertrand Moal MD, PhD, Taha Mohseni Ahooyi PhD, Jason H Moore PhD, Cinta Moraleda MD,PhD, Jeffrey S Morris, Michele Morris BA, Karyn L Moshal, Sajad Mousavi PhD, Danielle L Mowery PhD, Douglas A Murad, Shawn N Murphy MD, PhD, Thomas P Naughton BA, Carlos Tadeu Breda Neto, Antoine Neuraz MD, PhD, Jane Newburger MD, MPH, Kee Yuan Ngiam MBBS, FRCS, Wanjiku FM Njoroge MD, James B Norman, Jihad Obeid MD, FAMIA, Marina P Okoshi PhD, Karen L Olson PhD, Gilbert S. Omenn MD, PhD, Nina Orlova, Brian D Ostasiewski BS, Nathan P Palmer PhD, Nicolas Paris, Lav P Patel MS, Miguel Pedrera-Jiménez MS, Ashley C Pfaff MD, Emily R Pfaff PhD, Danielle Pillion MS, Sara Pizzimenti MS, Tanu Priya BS, Hans U Prokosch, Robson A Prudente PhD, Andrea Prunotto PhD, Víctor Quirós-González MS, Rachel B Ramoni, Maryna Raskin, Siegbert Rieg MD, Gustavo Roig-Domínguez MS, Pablo Rojo MD,PhD, Paula Rubio-Mayo MS, Paolo Sacchi MD, Carlos Sáez PhD, Elisa Salamanca, Malarkodi Jebathilagam Samayamuthu MD, L. Nelson Sanchez-Pinto MD, MBI, Arnaud Sandrin, Nandhini Santhanam MSc, Janaina C.C Santos MS, Fernando J Sanz Vidorreta, Maria Savino MS, Emily R Schriver MS, Petra Schubert MPH, Juergen Schuettler, Luigia Scudeller MD, MSc, Neil J Sebire MD, FRCPath, Pablo Serrano-Balazote MD,MS, Patricia Serre, Arnaud Serret-Larmande MD, Mohsin Shah MSc, Zahra Shakeri Hossein Abad PhD, Domenick Silvio, Piotr Sliz, Jiyeon Son MD, Charles Sonday, Andrew M South MD, MS, Francesca Sperotto MD, PhD, Anastasia Spiridou PhD, Zachary H. Strasser MD, Amelia LM Tan BSc, PhD, Bryce W.Q. Tan MBBS, Byorn W.L. Tan MBBS, Suzana E Tanni PhD, Deanne M Taylor PhD, Ana I Terriza-Torres MS, Valentina Tibollo MS, Patric Tippmann MSc, Emma MS Toh, Carlo Torti PhD, Enrico M Trecarichi PhD, Andrew K Vallejos, Gael Varoquaux, Margaret E Vella MPH, Guillaume Verdy MSc, Jill-Jênn Vie, Shyam Visweswaran MD, PhD, Michele Vitacca MD, PhD, Kavishwar B Wagholikar MBBS, PhD, Lemuel R Waitman, Xuan Wang PhD, Demian Wassermann, Griffin M Weber MD, PhD, Martin Wolkewitz PhD, Scott Wong, Zongqi Xia MD, PhD, Xin Xiong MS, Ye Ye BMED, MSPH, PhD, Nadir Yehya MD, MSCE, William Yuan PhD, Joany M Zachariasse MD, PhD, Janet J Zahner BS, Alberto Zambelli, Harrison G Zhang BA, Daniela Zöller PhD, Valentina Zuccaro MD, Chiara Zucco PhD
